# Cognitive Interview Validation of a Novel Household Hazard Vulnerability Assessment Instrument

**DOI:** 10.1101/2022.11.25.22281999

**Authors:** Taryn Amberson, Olive Ndayishimiye, Quanah Yellow Cloud, Jessica Castner

## Abstract

**Background:** Weather and climate disasters are responsible for over 13,000 USA deaths, worsened morbidity, and $1.7 trillion additional costs over the last 40 years with profound racial disparities. This project empirically generated items for a novel survey instrument of household hazard vulnerability with initial construct validation while addressing racial data bias.

**Methods:** Cognitive interviewing methodology was completed with transdisciplinary disaster expert panelists (n=20) from diverse USA regions on 60 unique hazard, disaster, or event items. Interview video recordings were qualitatively analyzed using thematic and pattern coding.

**Results:** A cognitive process mapped to themes of disaster characteristics, resources, individual life facet, and felt effect was revealed. 379 unique instances of linked terms as synonyms, co-occurring, compounding, or cascading events were identified. Potential for racial data bias was elucidated. Analysis of radiation exposure, trauma, criminal acts of intent items revealed participants may not interpret survey items with these terms as intended.

**Discussion:** The findings indicate the potential for racial data bias relative to water dam failure, evacuation, external flood, suspicious package/substance, and transportation failure. Hazard terms that were not interpreted as intended require further revision in the validation process of individual or household disaster vulnerability assessments.

**Conclusion:** Several commonalities in the cognitive process and mapping of disaster terms may be utilized in disaster and climate change research aimed at the individual and household unit of analysis.

**Highlights:**

⍰ Older adults and those with Black/African American racial identities are particularly susceptible to post-disaster health sequelae.
⍰ Prior to this study, no household-level Hazard Vulnerability Analysis existed. Quantifying risk for at-risk individuals/groups is a necessary initial step for working to eliminate disparities in large-scale disaster health outcomes.
⍰ Our findings indicate the potential for racial data bias relative to water dam failure, evacuation, external flood, suspicious package/substance, and transportation failure. Overall, several hazard, disaster, and event terms were not interpreted by survey-takers as intended, which may require elimination, replacement, or further revision in the validation process of individual or household assessments.

## Introduction

Climate change has been identified as the biggest threat facing humanity (1). Rising global surface temperatures increase the risk for and severity of droughts, floods, storms and other forms of severe weather and natural hazards (2). In the United States, weather and climate disasters are increasing in frequency and severity, responsible for over 13,000 deaths, $1.75 trillion additional costs, and worsened morbidity over the last 40 years (3-5). In 2021, the Centre for Research on the Epidemiology of Disaster (CRED) recorded an increase in the annual number of recorded global disasters (432 disasters in 2021 vs. previous 20 year annual average of 357) (6). No one is exempt from the adverse health risks of our changing climate, but some groups of people are disproportionately impacted by certain climate-sensitive health risks. These groups may also be more at-risk for negative and persistent health outcomes following disasters. Quantifying risk for specific individuals/groups is a necessary initial step in working to eliminate disparities in large-scale disaster health outcomes.

A hazard vulnerability analysis (HVA) is an assessment approach for identifying organizational level risks or hazards likely to impact certain facilities (often healthcare) and adjacent communities(7). These analyses enable a systematic risk calculation through the review of the probability of disaster events, institutional experience and capacity with disaster events, anticipated human, property, and financial impact with preparedness, internal, and external response planning(8). Institution-level HVAs are a mainstay to organizational-level and U.S. government disaster planning and mitigation efforts and are the basis of tailored planning and mitigation for risk reduction (9). Based on a systematic review of the literature, we identified that no household level HVA currently exists. The need for valid and reliable assessment of household HVA is necessary when considering individuals and households most vulnerable to the health effects of climate change and weather-related disasters. This work prioritizes people at the intersection of three at-risk groups: 1) older adults, 2) individuals with Black racial identities, and 3) those with chronic obstructive respiratory diseases (COPD, which includes chronic bronchitis, pulmonary emphysema, asthma. At this time, we have also included long-COVID with respiratory symptoms).

Older adults and those with Black/African American (B/AA) racial identities are particularly susceptible to respiratory symptoms, disease exacerbation, unscheduled health care utilization, and decreased quality of life after disaster exposure to particulates, mold, and flooding(3,10-14). Community-dwelling older adults with complex health needs are generally very poorly prepared for disasters (15-17), and account for half of recent disaster deaths (18). For older adults with COPD, disasters are linked to an increased risk for hospitalization in the 30 day window after the disaster (19). In the U.S., only 12% of households have the most basic elements of household disaster preparedness needed to shelter in place at home for 3 days(20-22). Further, profound racial disparities for those with Black racial identities have been observed in disasters such as the COVID-19 pandemic and weather-related disasters(23-26), exacerbated by long-standing disparities inherent to macro-level segregated housing and sociopolitical networks with fewer financial savings resources set aside for disasters (27-29). When controlling for pre-disaster disease burden, social network support, social vulnerability, and socioeconomic resources, racial disparity in other large-scale disasters was no longer associated with health outcomes like post-disaster depression(29-32). Given centuries-long structural racism with resulting segregated education, housing, and policy/law enforcement, multi-level conceptualizations and research designs are essential to adequate understanding of disaster-related racial disparity(33-36).

Self-reported data procedures on disasters, such as through survey instruments developed for the general population, may be particularly prone to validity problems and errors when utilized among historically marginalized or under-represented groups. Variations in how participants’ comprehension of the question wording, recall of information, meaning-making of their memories, and matching these ideas to the response options may result in very different information than the survey item developer had intended to obtain. Thus, the purpose of this research was to empirically generate items for a novel survey instrument of household HVA and initiate the process of validating (construct validity) these items using a process to minimize racial data bias. This research is the initial step of a multi-phase project (37) and focuses on the thematic and pattern coding and analyses of the 60 disaster/hazard terms (Figure 1).

**Figure 1.**
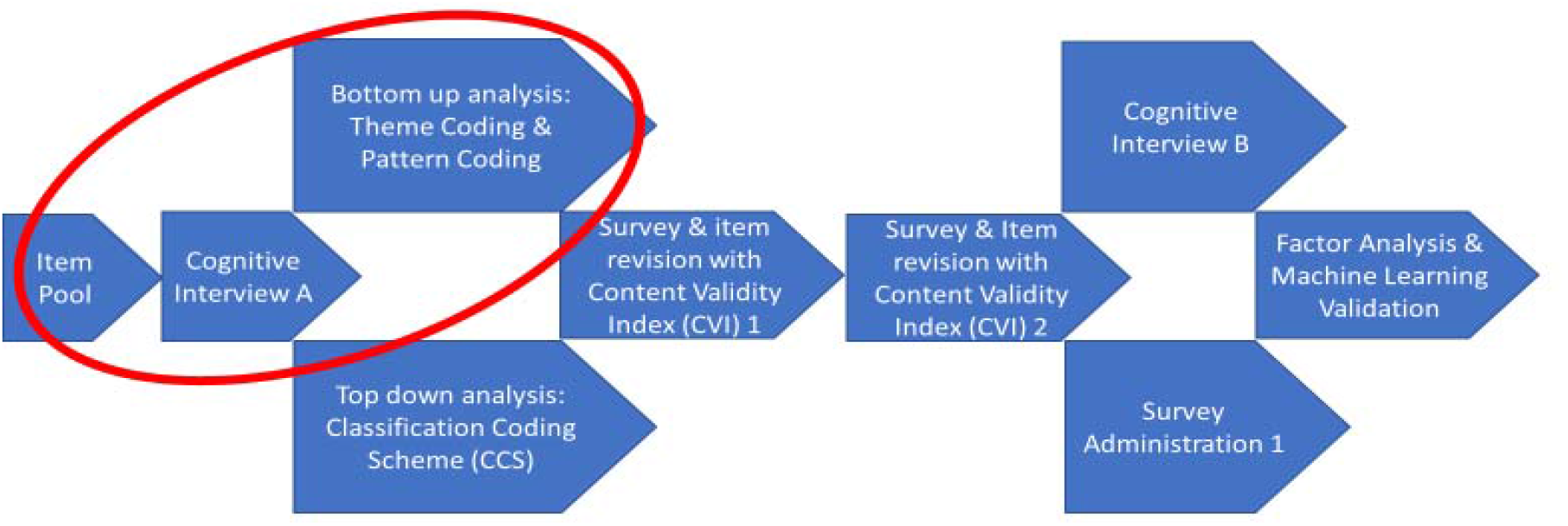
A depiction of how this manuscript fits into the overall project.

## Materials and Methods

### Qualitative approach and research paradigm

Cognitive Interviewing is a research method used to 1) identify problems participants may experience with survey questions, 2) study survey item construct validity, and 3) examine differences in thought processes in response to survey items across different demographic, linguistic, or cultural groups(38). Cognitive interviews facilitate understanding regarding the performance of the drafted survey questions; specifically, if the respondents understand the questions according to their intended design, and if accurate answers are given based on that intent. Utilizing this method with a diverse group of participants yields deep, contextual insight into how respondents interpret questions, consider relevant aspects of their lives and formulate responses based on those considerations(39).The technique can be used through a descriptive process for nascent survey items, or using a reparative approach to revise established items. Here, we utilized a descriptive process with an expert panel of 18 transdisciplinary participants, and a reparative approach with a final 2 additional participants.

### Sampling Strategy

Purposeful and snowball recruitment techniques were employed to assemble an expert panel of 20 members. We intended to sample a group of people who had a high likelihood of experiencing the disasters or hazards in the survey items we were testing. Thus, rather than future survey respondents, we sampled disaster experts at this stage due to the total number of disaster/hazard terms and the geographic variability in frequency of impact. Our strategy intentionally over-sampled (up to 50%) those with Black and/or African American racial, biracial or multiracial identities. See Appendix A for more information on our rationale for including 20 expert panelists.Inclusion criteria were a nationally or internationally recognized expert in their disaster-related discipline as evidenced by publications, awards, and/or fellowships and professional work experience as a first responder or disaster responder, public health, home health, emergency nursing, disaster nursing, and health care management expertise, and reside in the U.S. Our current national professional network enabled access to national experts through which we recruited. We recruited the first 18 participants for the full iteration of procedures, and a final 2 panel participants for a reparative approach after final item revision.

### Protection of Human Subjects and Data Security [Ethical issues pertaining to human subjects]

The protocol and study materials were reviewed by the Advarra Institutional Review Board (IRB) as application Pro00057555 and determined to be exempt from IRB oversight.

### Data Collection Methods, Instruments and Technologies

An item pool was created from existing organizational HVA’s (Kaiser Permanente, Risk Identification and Site Criticality Toolkit and CRED listings) and a literature review for use at the household level (9,40,41). A semi-structured cognitive interview guide was designed by a member of the research team (JC) with extensive experience in emergency nursing and instrument development and pilot tested with a consulting member of the expert panel.

Interviews were conducted by the same member of the research team (TA) from December 2021 through May 2022. The purpose of the interviews was to ascertain perceptions of standard HVA domain items and responses(8) when applied to the household level. Refer to Appendix A for additional detail regarding the interview structure.

Demographic information collected included sex at birth, current gender, age, racial and ethnic identities, language spoken in the home, veteran status, household member veteran status, and highest completed level of education

### Data Processing and Analysis

The videorecording of the interview was utilized as the raw data for analysis. No transcription was used in order to fully incorporate non-verbal information in the analytic process(42,43). Interviewer notes augmented the videorecording data. Theme and pattern coding were used for analyses (Figure 1). Refer to Appendix A for additional detail regarding data analyses.

### Techniques to Enhance Trustworthiness

We maintained an audit trail and triangulation to enhance trustworthiness and credibility of data analysis. Our audit trail includes original recordings, double-entered interviewer notes, double-reviewed interviews for thematic codes, and duplicate files for each stage. The findings were triangulated with a content validity index, which will be reported elsewhere. Member checking is ongoing as part of the multi-faceted, overarching project.

### Context

All participants confirmed they were located within the United States of America (USA) or USA territories at the time of the interview. The interviews were conducted over the web and recorded (audio and video, as available). We allotted 90-minutes for each meeting with expert panelists. Here, we report the results of our thematic and pattern coding analysis(44).

## Results

### Expert Panelist Characteristics [Units of Study]

This transdisciplinary panel represented a variety of occupations, including epidemiology, chemistry, fire service, first responder, nursing, academic professor, consulting and roles of disaster planning and response throughout all phases of the disaster management cycle. Panelists’ areas of professional expertise reflected disaster-related leadership, expertise and service that spanned all levels of government (local/state/national/international) and included (but were not limited to) sectors of public health, emergency preparedness, management and response, emergency medical services, global health security, non-profit engagement and health care leadership.

Table 1 lists the demographic characteristics of the expert panel. Of the initial 18 interviewers completed with a descriptive approach, seven (38.8%) expert panelists identified their current gender as “man,” and their sex as male. Ten (55.5%) expert panelists identified their current gender as “woman” and their sex as female, and one respondent chose not to report their sex or gender. Eight (44.4%) expert panelists were between the ages of 30-49, and five (27.7%) were between the ages of 50-64, and another five (27.7%) were 65 and older. Eight expert panelists (44.4%) endorsed a B/AA racial identity. Of these, 2 participants indicated additional racial identities (Biracial, White/Caucasian, Native American, or a combination) to B/AA. Eight expert panelists (44.4%) endorsed a Caucasian racial identity. Of these, 1 participant indicated an additional racial identity of Native American in addition to White/Caucasian. One expert panelist endorsed a Native Hawaiian/Other Pacific Islander racial identity, and the final expert panelist interviewed with a descriptive approach vocalized the desire for an “Other” or blank/fill-in category in response to this question. All participants spoke English, with two participants also speaking other languages in their home (Arabic and French). Collectively, panelists mapped their answers pertaining to personal or professional experience of the 60 hazards/disasters to every region of the Mainland USA, Alaska, Hawai’i, Puerto Rico and the Virgin Islands. International geographies mentioned as participants cognitively mapped their experience with each hazard or disaster term included Japan, Afghanistan, Honduras, Haiti, West Africa and China. The demographics of the two expert panelists interviewed with a reparative approach are included in Table 1 as well.

**Table 1.** Demographic Characteristics

| Table 1 | Descriptive Approach | Reparative Approach |
| --- | --- | --- |
| Demographic Characteristics | n = 18<br>n (%) | n = 2<br>n (%) |
| Sex at birth |  |  |
| Male | 7 (38.9%) | 1 (50%) |
| Female | 10 (55.6%) | 1 (50%) |
| Refused | 1 (5.6%) | 0 |
| Current gender |  |  |
| Man | 7 (38.9%) | 1 (50%) |
| Woman | 10 (55.6%) | 1 (50%) |
| Other | 1 (5.6%) | 0 |
| Age, year |  |  |
| 30-49 | 8 (44.4%) | 1 (50%) |
| 50-64 | 5 (27.8%) | 1 (50%) |
| 65 and Over | 5 (27.8%) | 0 |
| Marital status |  |  |
| Married couple | 13 (72.2%) | 2 (100%) |
| Separated/divorced | 3 (16.7%) | 0 |
| Never married | 2 (11.1%) | 0 |
| Veteran status – Have you ever served in the U.S. military? |  |  |
| No | 13 (72.2%) | 2 (100%) |
| Active duty | 1 (5.6%) | 0 |
| Reserve or National Guard | 2 (11.1%) | 0 |
| Both Active Duty and Reserve or National Guard | 2 (11.1%) | 0 |
| Veteran status – Have members of your current household ever served in the U.S. military? |  |  |
| No | 16 (88.9%) | 1 (50%) |
| Active duty | 2 (11.1%) | 1 (50%) |
| Reserve or National Guard | - | - |
| Both Active Duty and Reserve or National Guard | - | - |
| Main household language |  |  |
| English | 16 (88.9%) | 2 (100%) |
| English and other (French, Arabic) | 2 (11.1%) | 0 |
| Racial identities* |  |  |
| American Indian/Alaskan Native | 1 (5.6%) | 0 |
| Asian | - | - |
| Black or African American | 8 (44.4%) | 1 (50%) |
| White/Caucasian | 8 (44.4%) | 1 (50%) |
| Native Hawaiian or Other Pacific Islander | 1 (5.6%) | 0 |
| Don't Know/Unsure ("Other"**) | 1 (5.6%) | 0 |
| Refused |  |  |
| Ethnic identity<br>Hispanic, Latinx or of Spanish origin (Yes) | 0 | 0 |
| Education level |  |  |
| Less than high school completion/diploma | - | - |
| High school degree/GED/or equivalent | - | - |
| Some college, no degree | 0 | 1 (50%) |
| Associate's degree | 1 (5.6%) | 0 |
| Bachelor's degree | 0 | 1 (50%) |
| Master's, doctorate, or professional degree | 17 (94.4%) | 0 |
| Table footnotes: *Participants could choose more than one category for racial identity; Percentages may not add up to 100% due to rounding; **Category of "Other" was requested by the participant |  |  |

### Pattern Coding Results

The results of the pattern coding we utilized are depicted on Table 2. Here, we used pattern coding to assess for the potential for racial data bias and ascertain group differences between those with B/AA racial identities and those who did not report any B/AA racial identity.

**Table 2:**
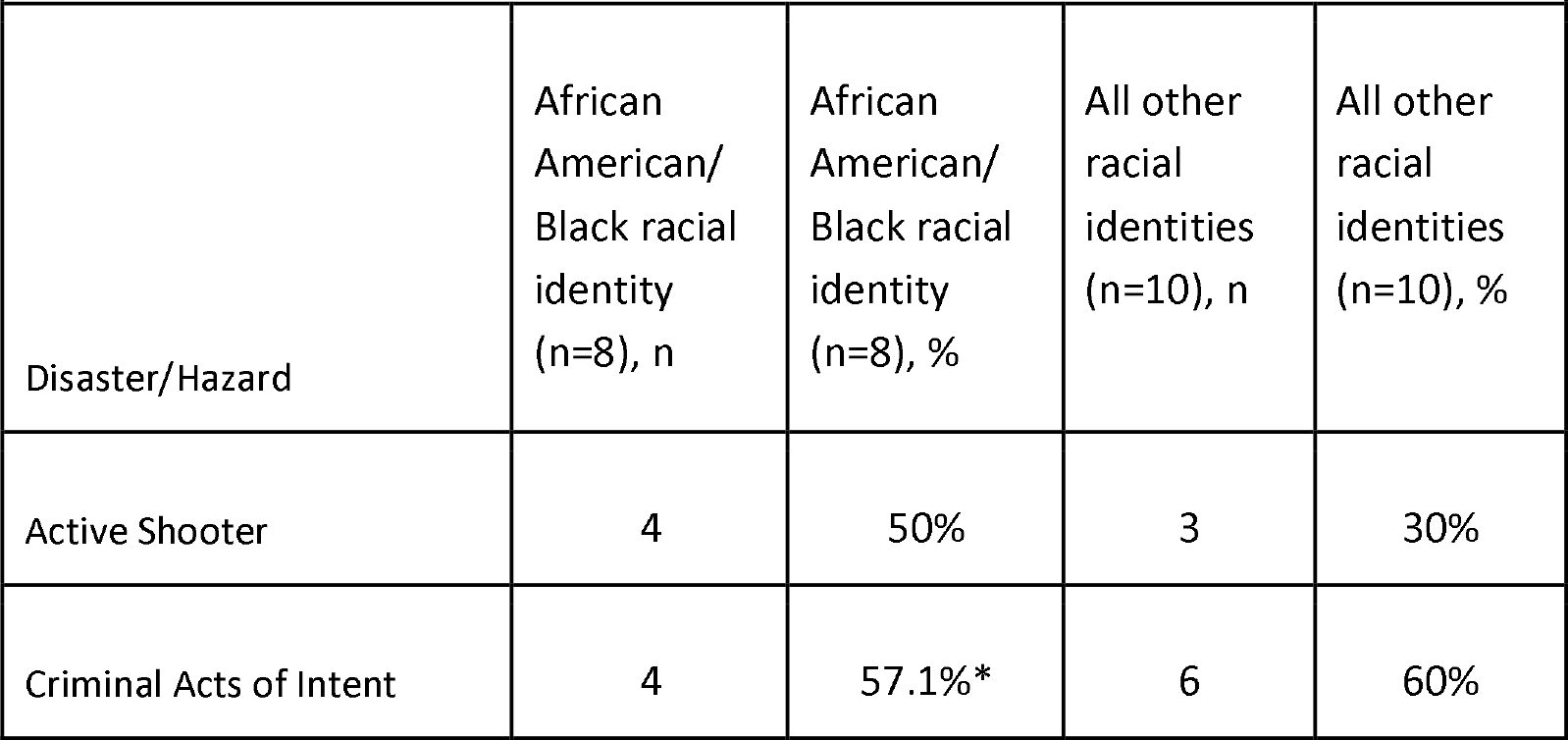
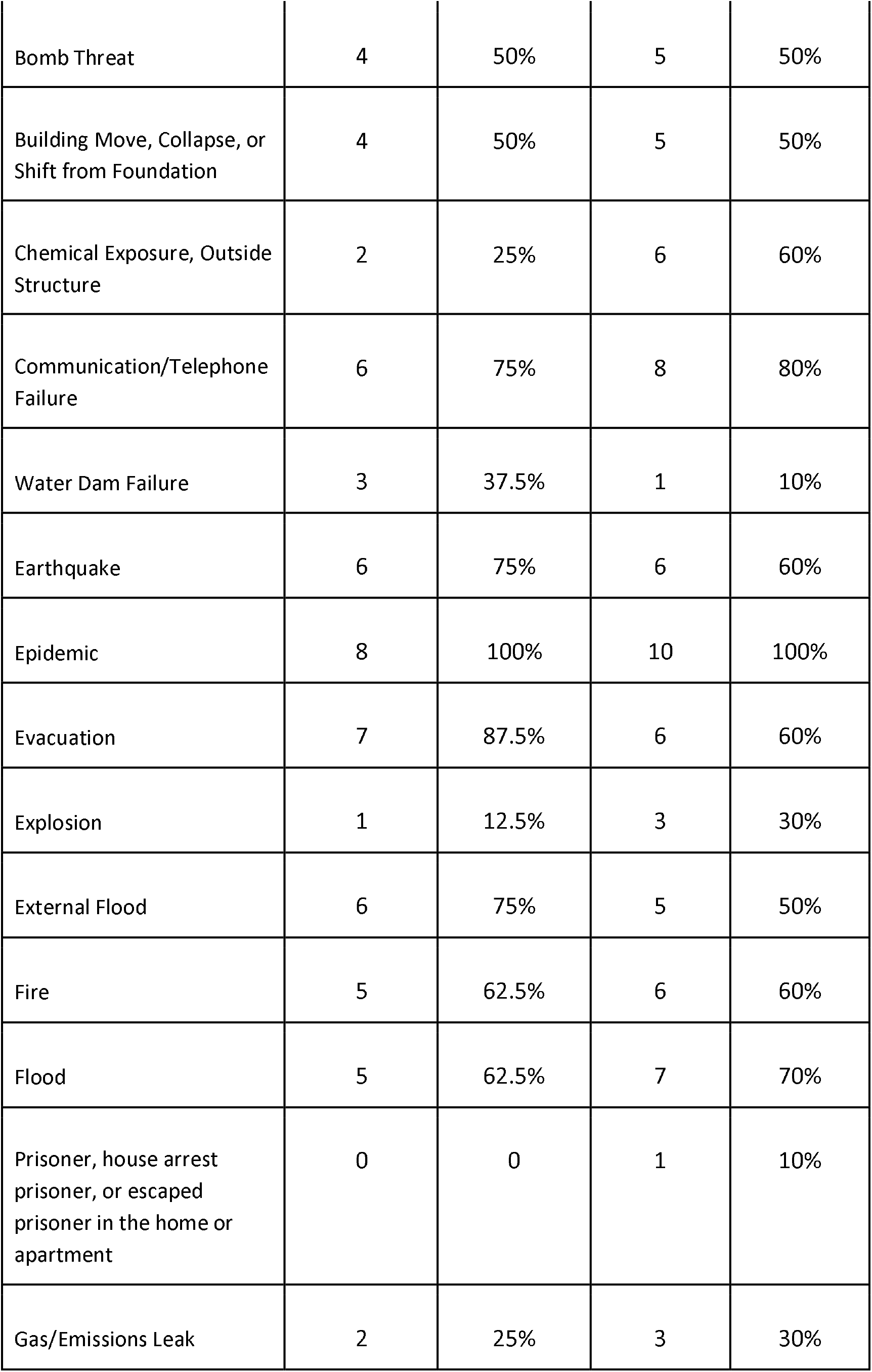
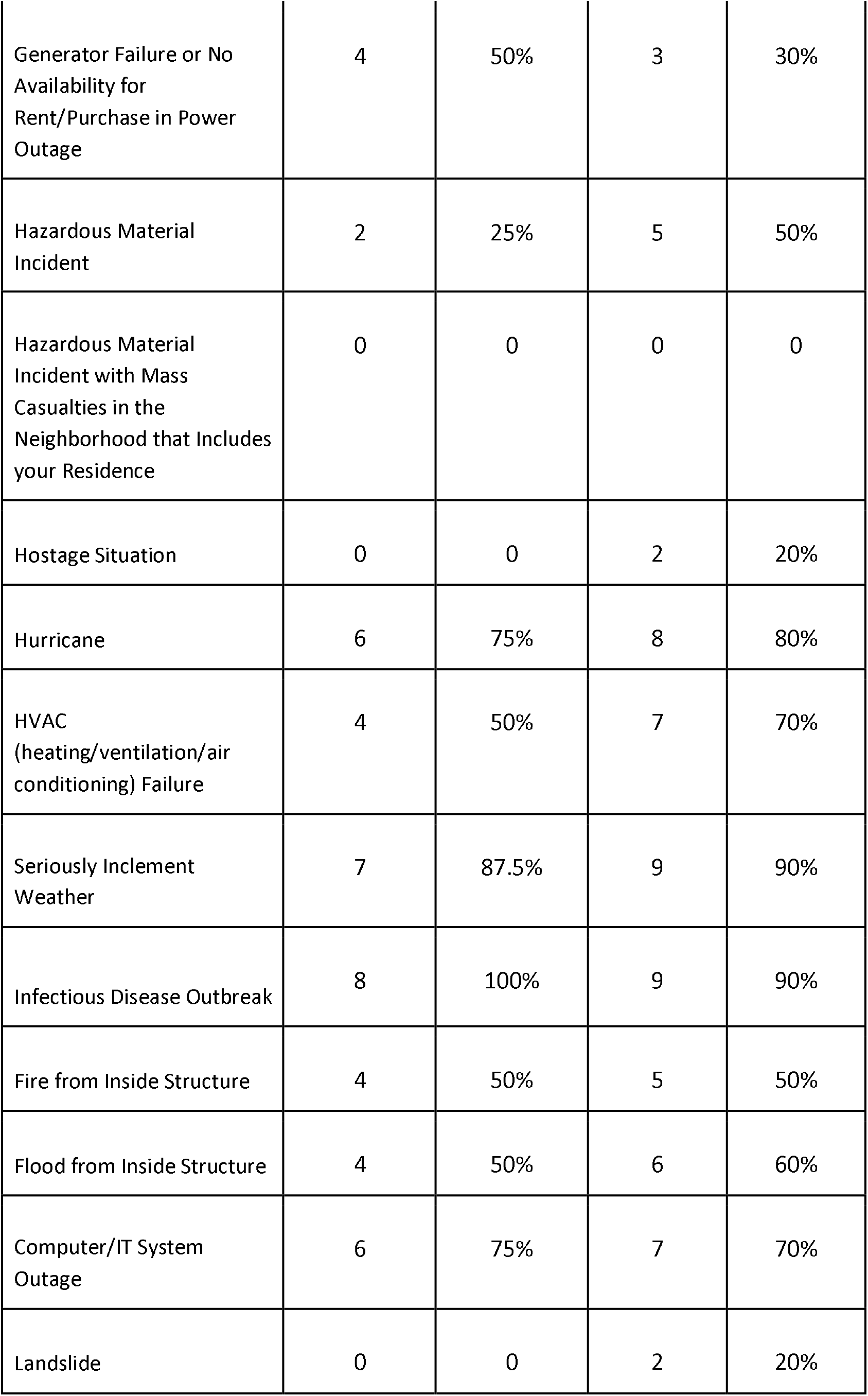
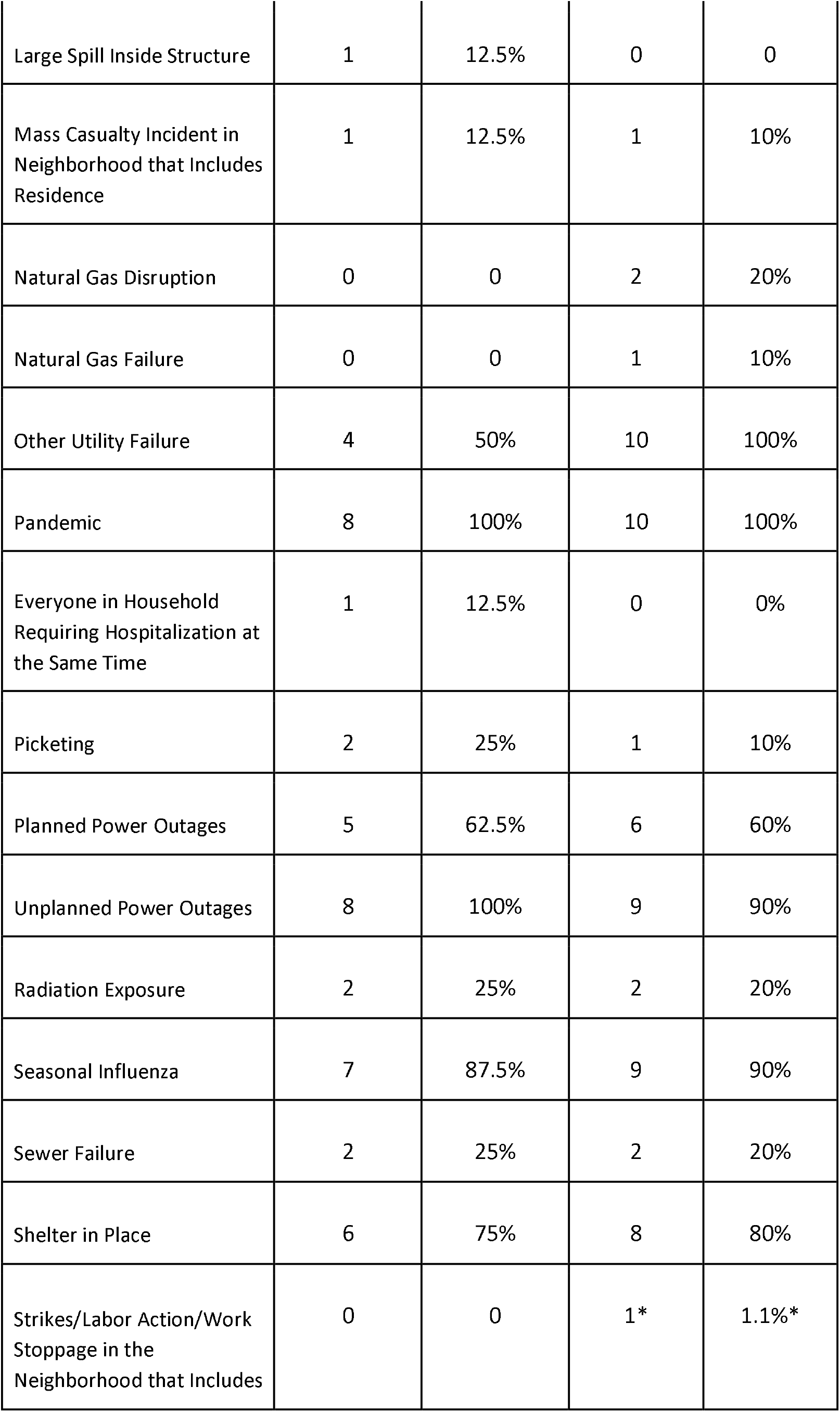
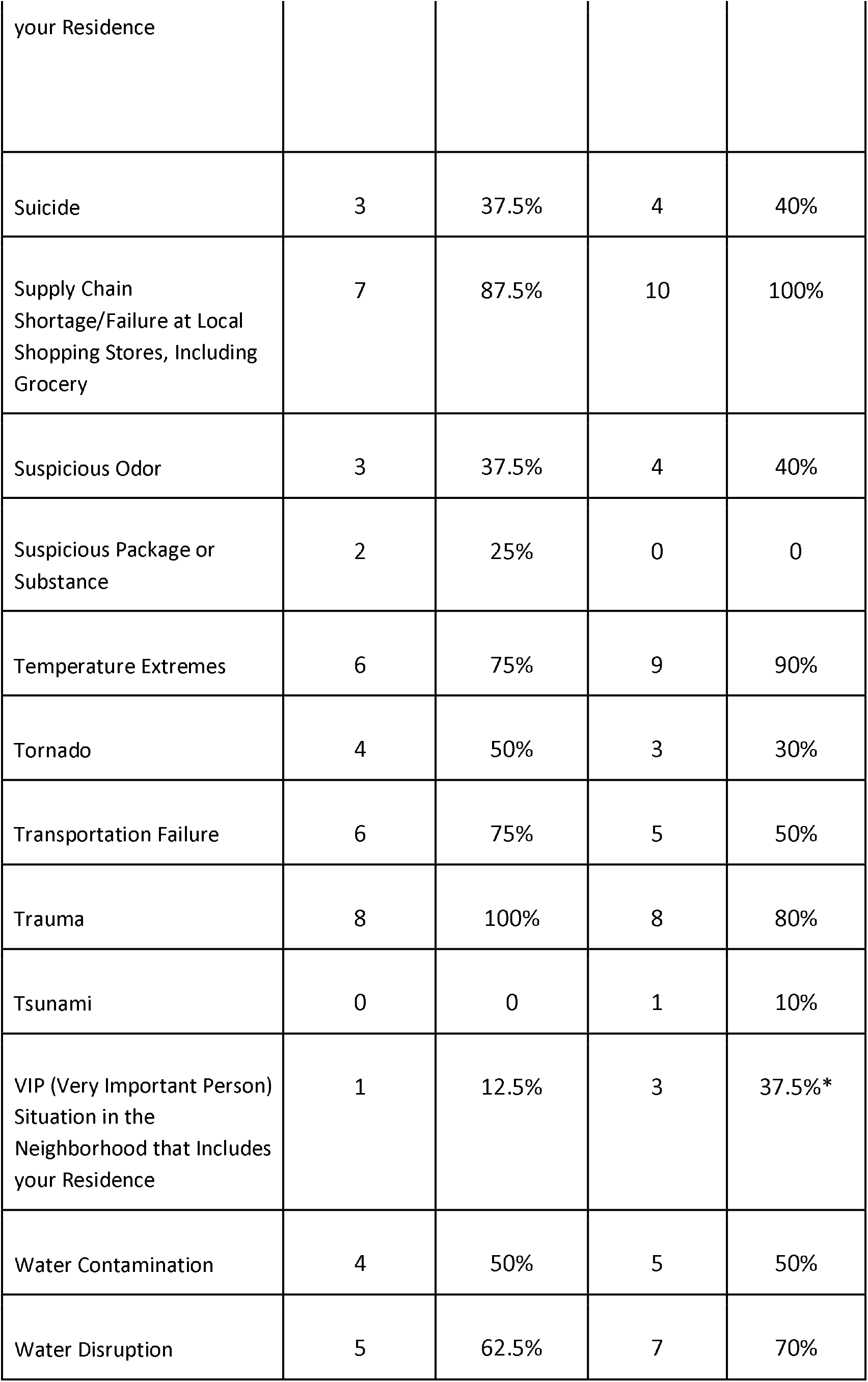
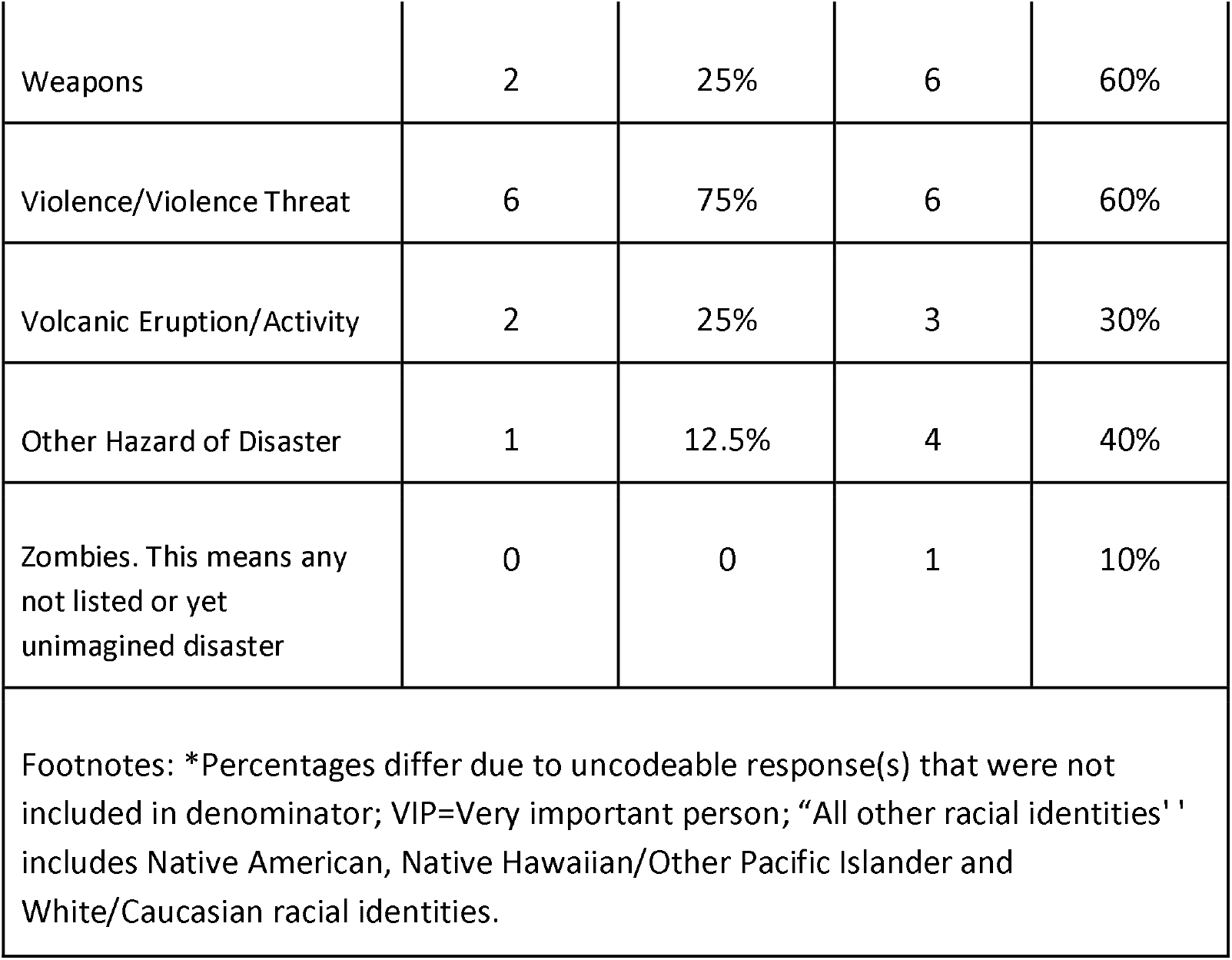
Pattern coding results Frequency (n and %) of “Yes” to Direct Impact by Disaster/Hazard Impact, n=18 interviews

### Theme Coding Results

Overall, across participants and items, four overarching themes emerged in the general conceptualizations examined in response to the items worded as, “In your lifetime, have you ever been directly impacted by [disaster/hazard/event term]?” (See Appendix B for an example). These themes were the disaster characteristics, resources, individual life facet, and felt effect. Figure 2 depicts a flowchart of these themes with subthemes. Participants consistently mapped their stories and responses along this overarching flowchart as part of both their comprehension of the question and in judging or justifying their retrieved information as warranting the response as either yes or no. For example, one participant endorsed a secondary, emotional impact to the active shooter disaster/hazard. Although they were not physically present for the actual event, they felt emotionally impacted, although “not directly,” while supporting family members (including minor children) who were processing and psychologically recovering following this event. In addition, participant’s cognitive walk-through of individual life facets was also used as an aid in the retrieval of relevant information from memory.

**Figure 2.**
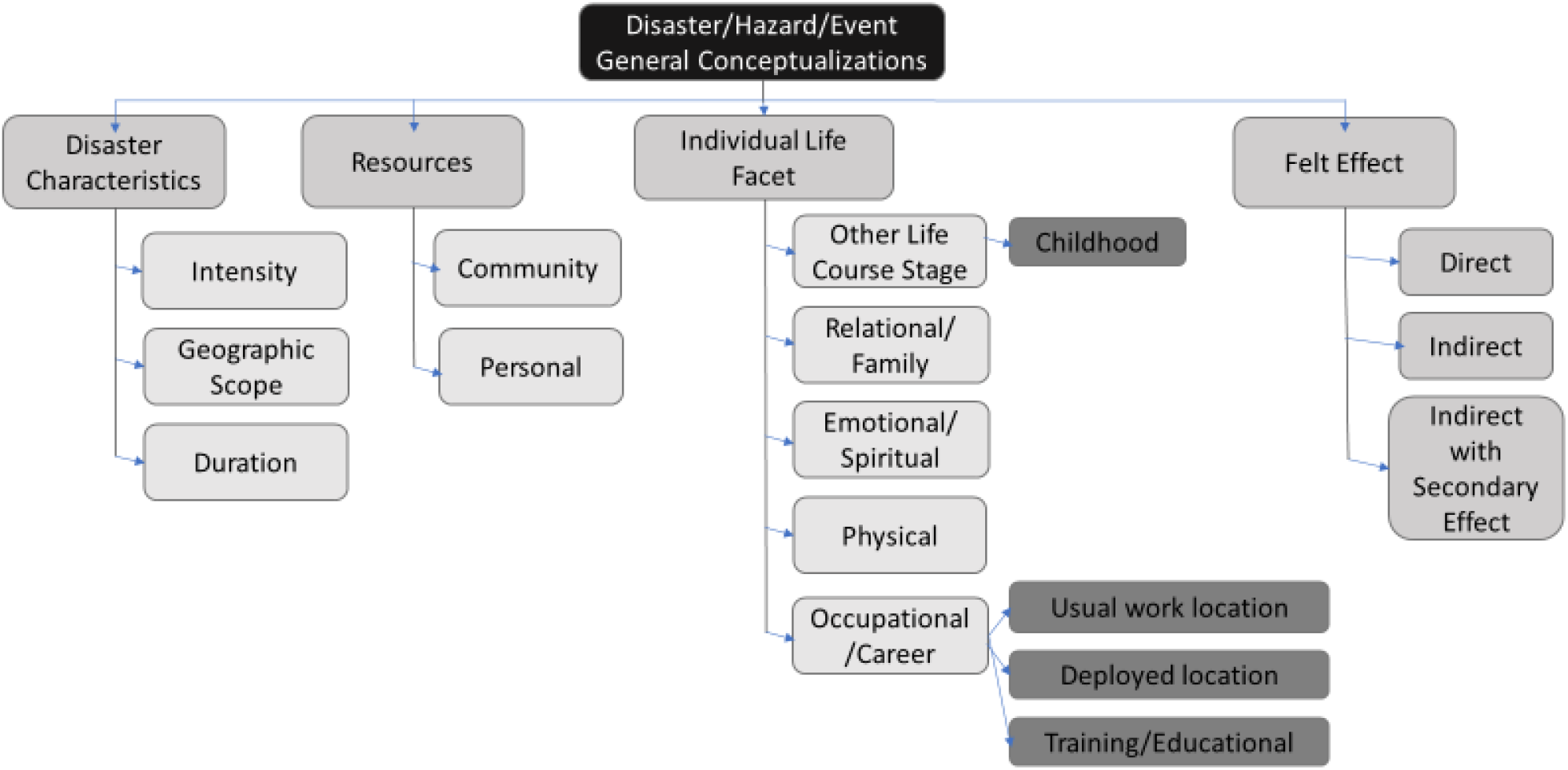
Flowchart organization of themes uncovered across all items and participants

#### Linked terms

The theme coding process was also used to generate item-specific schemas for each of the 60 hazard/disaster/event terms, revealing variation and commonalities across participants in their understanding of each term or concept. Across many of the terms, the question-answer narrative revealed multiple synonyms with other terms, co-occurring, compounding, or cascading events with other survey item terms. For example, across the 18 expert panelists interviewed with a descriptive approach, the term water disruption was compounded with other disasters/hazards like other utility failure, water contamination, sewer failure, hurricane, temperature extremes, seriously inclement weather and unplanned power outage. In another example, the term external flood revealed synonyms, co-occurring and cascading events with other survey items terms like hurricane, flood, seriously inclement weather, evacuation, water contamination, internal flood, flood, telephone/communication failure/disruption and water dam failure. One participant stated when they were a child, their “house was under a boil water order, but the flooding did not actually impact where I was staying, but I relocated to a shelter as part of the flood.” This participant recalled disruption to telephones as a cascading event. We coded a total of 379 unique instances of linked terms across the 18 interviews and 60 terms. As an example, figure 3 provides a data visualization of the 20 instances of terms linked to “Building Move, Collapse, or Shift from Foundation” in a Chord Diagram.

**Figure 3.**
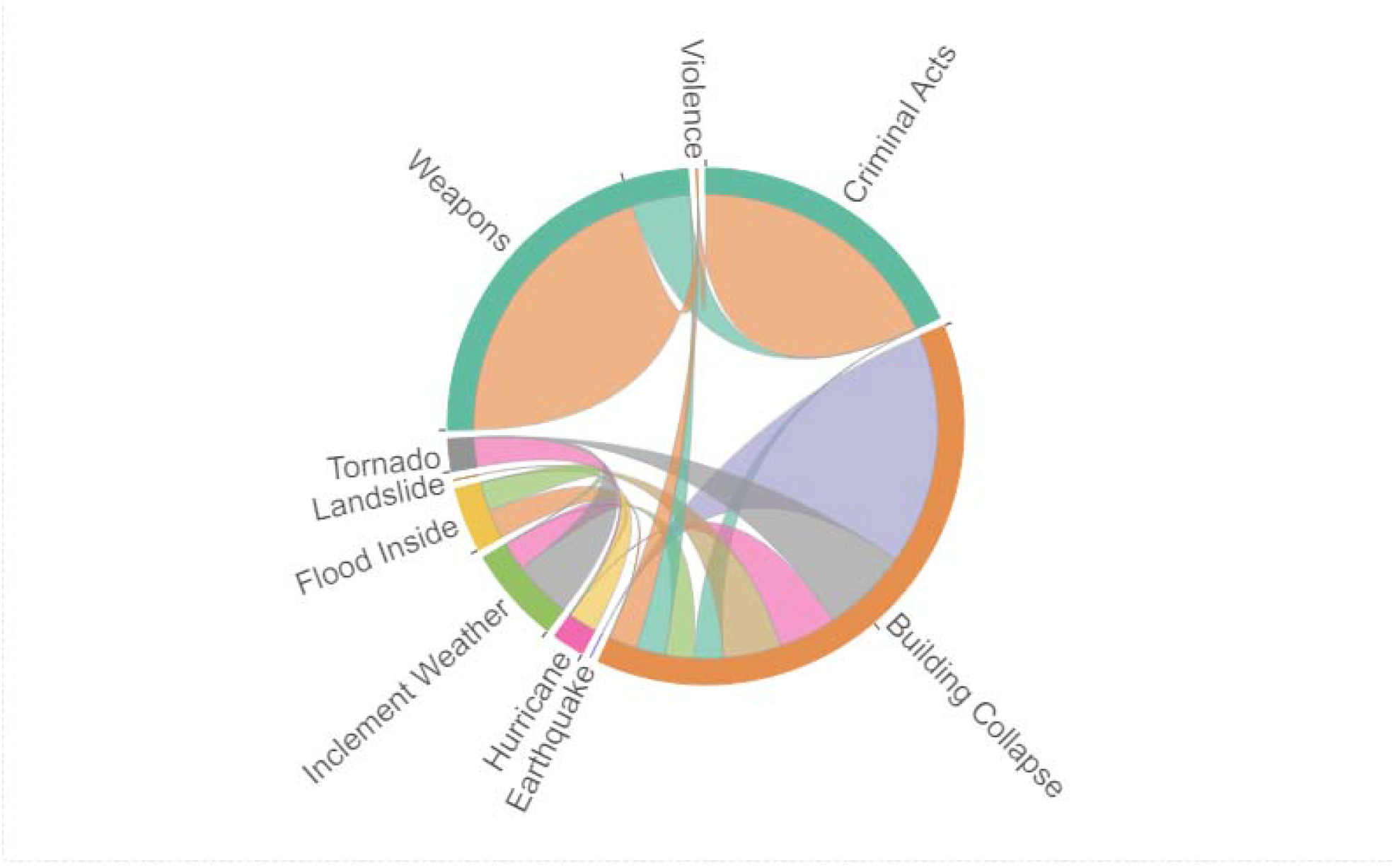
Figure 3. Chord Diagram of terms linked to Building Move, Collapse, or Shift from Foundation Figure 3 note: Building Move, Collapse, or Shift from Foundation was linked to other terms a total of 20 times: Earthquake (8); Tornado (3); Hurricane (2); Seriously Inclement Weather (1); Landslide (2); Criminal Acts of Intent (1); Weapons (1); Violence (1), Flood Inside Structure (1). Several of these terms were also linked to one another (e.g. Seriously Inclement Weather and Tornado).

#### Item Specific Schemas

Item specific schemas emerged from the theme coding as well. As an example, the item-specific schema for water contamination revealed additional detail to the general flowchart the mapped common cognitive processes as participant’s narratives focused on details relative to the antecedents of the water contamination, physical location within various life facets, and specific consequences and felt effects. Figure 4 details these additions to the general schema elucidated from the theme codes.

**Figure 4.**
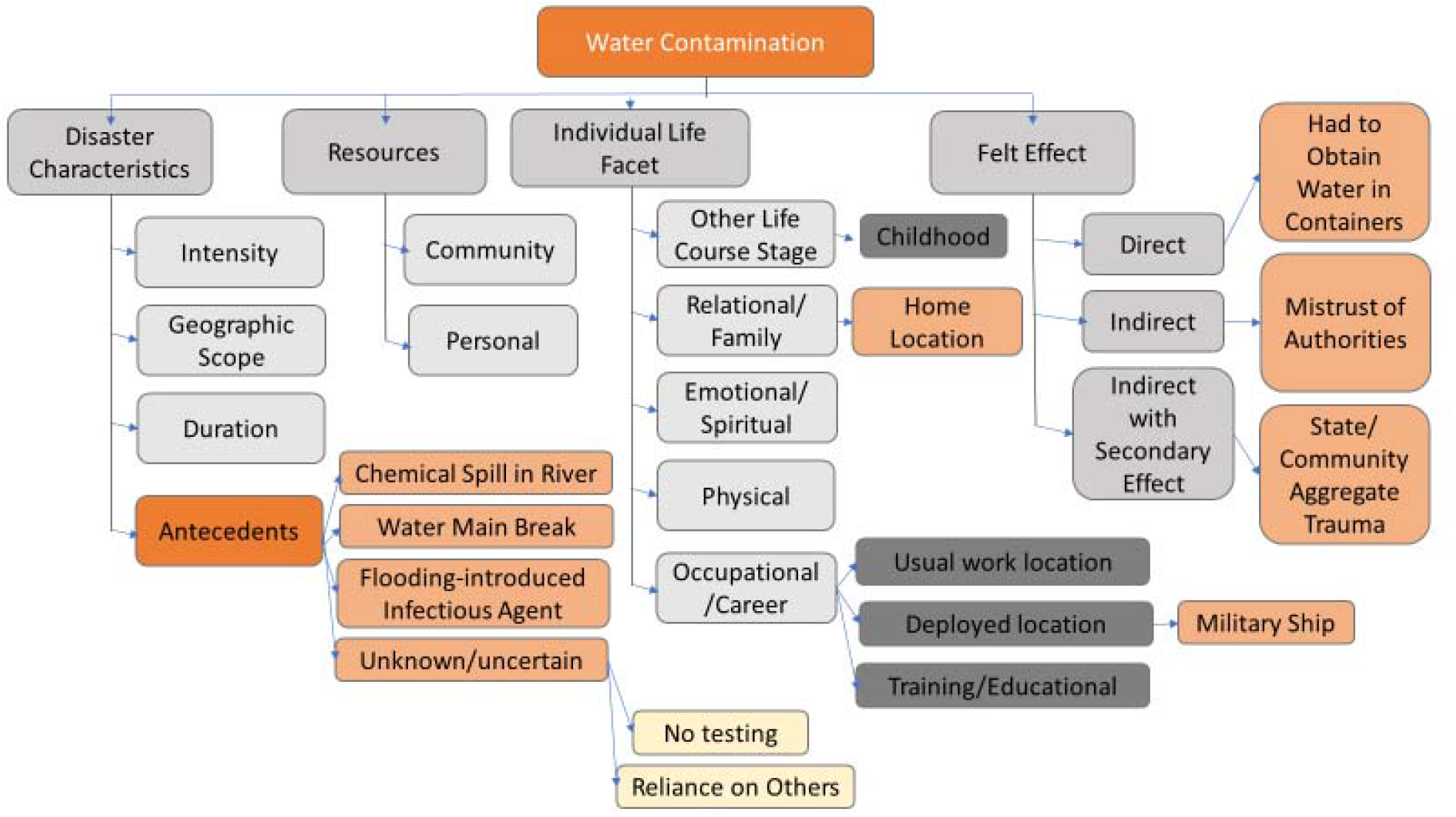
Schema of themes specific to Water Contamination across participants

Other disaster terms elucidated specific major events or specific places in the recall and cognitive mapping of the term. For example, the term hurricane evoked responses and memory retrieval specific to unique disaster events, such as Hurricane Katrina or Hurricane Ida (See Figure 5). Alternately, the term Active Shooter evoked narratives and memory retrieval specific to the location of the event (see Figure 6).

**Figure 5.**
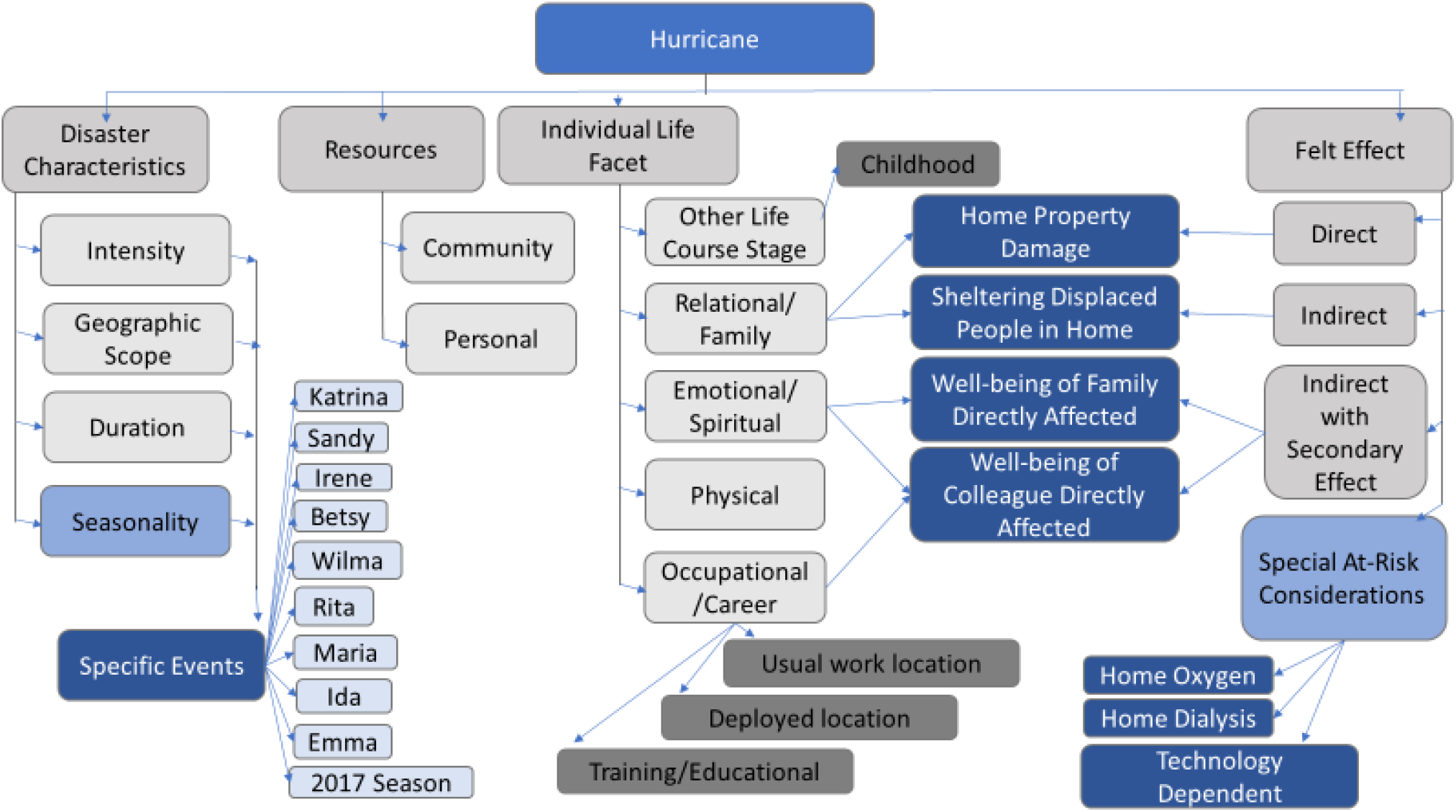
Schema of themes specific to Hurricane across participants

**Figure 6.**
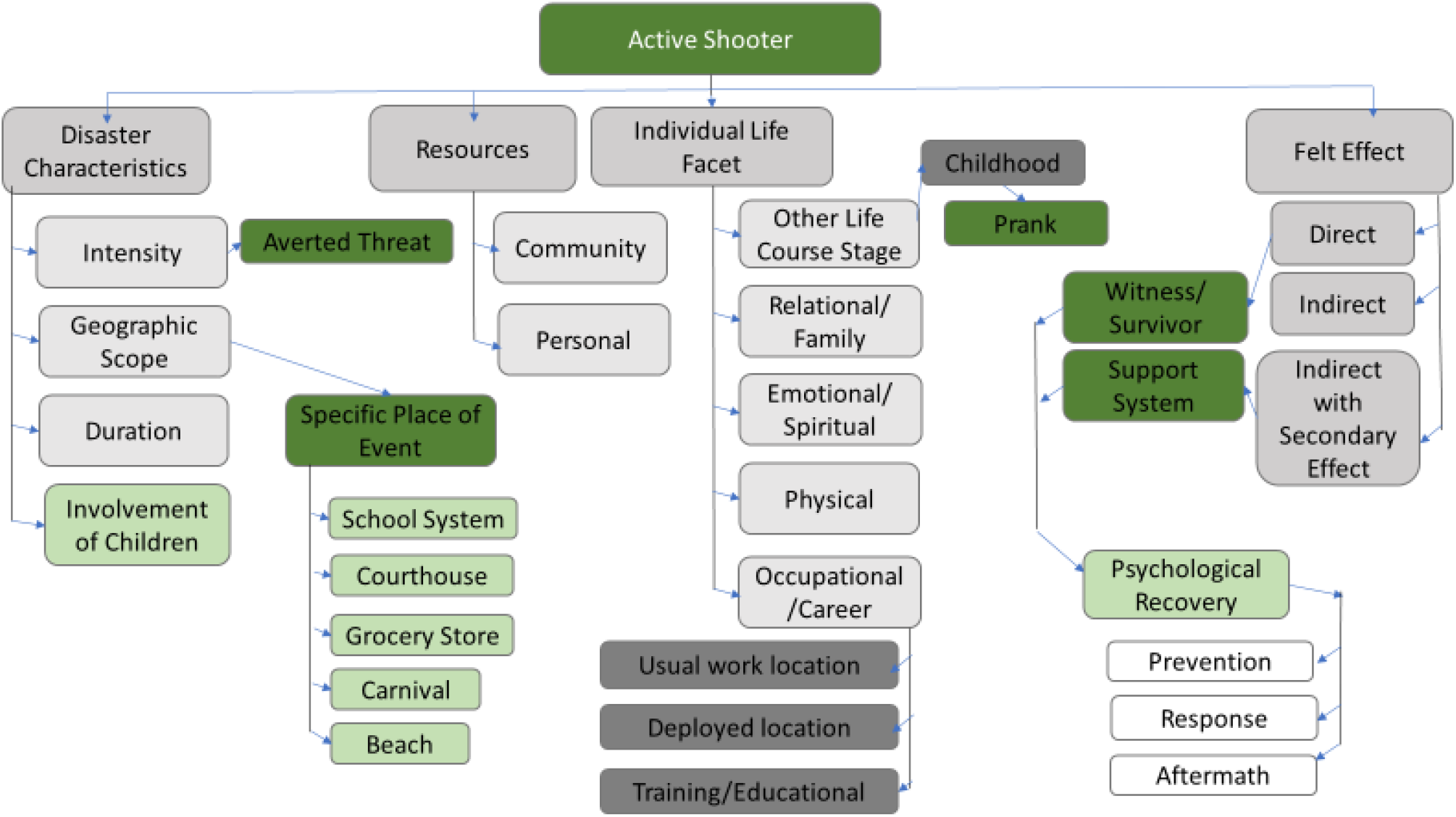
Schema of themes specific to Active Shooter across participants

### Synthesis of Results by Problematic Terms

Of the 60 disaster terms, the cognitive interviewing and analysis process revealed potential problems with several of the disaster, hazard, and response terms. These terms require elimination, replacement, or further revision in our validation process and survey instrument development. In Appendix B, we summarize the findings of the three terms we identified as most problematic or with the most disparate interpretations by participants. These terms were trauma, radiation exposure, and criminal acts of intent. The findings specific to these terms are summarized below, integrating a synthesis of the empirical data from direct interview quotes, field notes and theme coding.

#### “Trauma”

When asked if they’ve ever been directly impacted by “Trauma” in their lifetime, multiple respondents asked if this question referred to physical or psychological trauma; with some answering this item only thinking of one form or the other. At least 5 respondents identified this as a complex or vague topic. One respondent replied, “Trauma, that’s a big word,” and another identified it as a “popular” word. At least 7 participants exhibited a long recall period or required high detail, not always being able to access the information through recall. This term was also flagged as being potentially sensitive or prone to desirability bias in at least 3 interviews. At least 6 respondents requested clarification or expressed uncertainty regarding this term.

This question captured several themes across respondents, including primary physical trauma (car crash; fall, gun shot wound, crush injury; childbirth; near-death injury/illness) and psychological trauma (COVID 19 sequelae, coping with friend’s suicide, LGBTQ+). However, psychological trauma was not independent of physical trauma in panelists’ answers to this item. Vicarious, relational trauma through witnessing the suffering, illness and/or death of loved ones and vicarious trauma from occupational experiences were also mentioned.

Across groups, 100% (n=8) of participants with B/AA identities (inclusive of multi and biracial Black identities) said they had been directly impacted by trauma in their lifetime. One respondent asked, “…Who among us has not been impacted by trauma?” This same respondent also said, “Everyone has been touched by trauma. So that may be one where you need to clarify a bit or give a timeframe.” Among participants of other racial identities, 80% (n=8) said they had been directly impacted by trauma in their lifetime. One respondent hesitated before answering “no.” When probed about what the term meant to them, they explained, “For me, when I think of trauma I think of physical trauma…there’s a broader term now…to include, or to frame, psychological trauma.” The respondent chose to answer this question only considering physical trauma, or “the physical environment,” to maintain consistency with how they answered other questions. The other respondent that answered this question as “no” considered close secondary impact from a family member’s traumatic experience.

#### “Radiation Exposure”

Some terms reflected diversity in Expert Panelists’ perceptions; influenced by variability in personal and occupational experiences, identities and geographical locations. The disaster/hazard term “Radiation Exposure,” for example, was mapped by respondents to ideas like background exposure (sunlight), medical treatment (cancer) and diagnostic procedures (X ray), occupational exposure, intentional release of radioactive material, transportation accidents with radioactive material, and power plant emission issues.

Some respondents exhibited difficulty trying to source and determine the scope of impact when answering this question. For example, one respondent initially answered “I don’t think so,” and thought out loud about being aware or unaware of healthy versus unhealthy levels and sources of radiation exposures; personally or occupationally. Multiple respondents mentioned routine exposures to radiation through ultraviolet rays, radon from basements and diagnostic procedures. One respondent considered where they lived in relation to nuclear power plants.

Across respondents, interpretations of radiation exposure were categorized as beneficial versus harmful (some overlap was noted with criminal acts of intent), by occupation, by event, and unknown exposures (i.e., missing radioactive materials, radioactive dispersal devices, contamination). This question also captured historical radiation-related disasters like Chernobyl and 3-Mile Island.

Across groups, 25% (n=2) of participants with B/AA identities (inclusive of multi and biracial Black identities) said they had been directly impacted by radiation in their lifetime. These two participants considered occupational and background/environmental sources of radiation exposure when answering this question. Among participants of other racial identities, 20% (n=2) said they had been directly impacted by radiation exposure in their lifetime. These two participants answered this question when thinking of health related (diagnostic or curative) exposures.

#### “Criminal Acts of Intent”

Examples of responses to the disaster/hazard term “Criminal Acts of Intent” included “What exactly is that?,” “I don’t know, that could be anything…,” “It just seems so broad,” and “That is a mouthful of a word…I think the answer is probably yes, but…I can’t be specific.” Respondents mentioned instances of theft, breaking and entering, assault or other acts with intent to hurt or extremely inconvenience a group of people, system or setting. One expert panelist used terms like espionage, ransomware or drugs and another described incidents of workplace violence and verbal aggression in public spaces. At times, participants answered “yes” to this question, but could not map this term to a specific memory or access the information through recall.

This term seemed to generate confusion among expert panelists. One respondent provided an uncodable answer of “I don’t know.” Eight respondents expressed uncertainty and/or requested clarification of the term, with two of these also requiring a repeat of the question. This item was identified as a vague topic/term by two participants.

Across respondents, patterns of interpretation included state level - terrorism/espionage; household level theft (car or home); breaking & entering; personal or interpersonal group level aggressive behaviors (verbal and non-verbal) with implicit or explicit threats of violence; and adolescent misconduct (implications of non-violent pranks to gang behaviors).

Across groups, 57.1% (n=4) of participants with B/AA identities (inclusive of multi and biracial Black identities) said they had been directly impacted by criminal acts of intent in their lifetime. One respondent with a B/AA racial identity provided an uncodable answer, but indicated in their answers to interviewer probes they have experienced this term in similar ways to how other panelists interpreted it. Among participants of other racial identities, 60% (n=6) said they had been directly impacted by criminal acts of intent in their lifetime. One participant noted “Having things stolen was not a disaster,” which became an emerging theme throughout the interviews with expert panelists.

## Discussion

Here, we have reported the results of empirically generating and validating items for a novel survey instrument of household HVA using a cognitive interviewing process to minimize racial data bias. To the best of our knowledge, our work is the first to identify instrument development specific to the household disaster preparedness of older adults with COPD with a focus on the increased risk to those with B/AA racial identities. Our work contributes uniquely to the literature by 1) mapping a common cognitive processes in response to items with disaster, hazard, or disaster response terms, 2) mapping additions to this cognitive process for specific disaster contexts, 3) elucidating synonym, co-occurring, and compounding disaster events, and 4) detailing cognitive difficulties with particularly problematic or vague disaster terms. Generating a novel application and instrument using the cognitive interviewing process, there is little existing literature to which to compare our specific results and use of the methodology(45). Currently disaster related household assessments quantify current household preparedness status, high risk functional or health conditions of household members, or rapid needs assessment in the midst of a disaster(46,47). Our focus was on developing and validating an instrument relevant to disaster experience and measuring how this experience informed future household disaster risk and vulnerability. This work is timely and important to inform public health and clinical climate change vulnerability assessments and disaster planning (48). Our work is also crucial to developing climate change equity interventions focused on health and education(49), such as enhancing precision in the direct provision of disaster preparedness supplies or household disaster planning services to those identified as most at-risk or affected.

### Racial Identity

As identified in our pattern coding results (Table 2), we noted several areas that flag the need for further investigation into possible racial disaster disparities. We found greater than 30% difference, with a greater proportion of participants with all other racial identities reporting experiences with chemical exposure outside structures, other utility failure, and weapons, compared to participants with B/AA racial identities. These items were considered for elimination and/or combination with other terms for an instrument relevant to those with B/AA racial identities. We found a difference greater than or equal to 25% for water dam failure, evacuation, external flood, suspicious package/substance, and transportation failure with a greater proportion of those with B/AA racial identities reporting an experience with these events, compared to others. In contrast, a greater proportion of those with other racial identities reported experiences with explosion, hazardous material incident, very important person (VIP) situation, or other hazard than participants with B/AA racial identities.

Our findings align with previously published literature on the impact of historical structural racism for communities of color increasing the risks of negative health impacts related to flooding, transportation, and evacuation(23-26). Segregated neighborhoods and social networks generate racial disparities on a macro level (42,43) and perpetuate increased disaster-related racial disparities. Residents in low-income racially segregated communities, disproportionately overrepresented by Black residents, experience higher incidents of disaster toxic exposure(11), and worsened housing and resource recovery after disaster(51). Some people with B/AA racial identities may both reside in high risk disaster areas and have the least amount of resources to protect themselves and their families against or recover from climate change related hazards or disasters (50). Given the longstanding history of structural racism and unequal race-related wealth distribution in the United States, multi-level conceptualizations and research designs are required to better understand disaster-related racial disparity(39-42), especially when collectively investigating climate change, health equity and household-level disaster preparedness.

In addition to no household-level hazard vulnerability assessment, our team found no cognitive interview reports online through QBank in the last 5 yeast that investigate data or racial bias for individuals or communities with B/AA racial identities(45,51). In the context of health disparity research with substance abuse, Burlew et al (2009) notes the importance of adequate measurement and cultural equivalence/appropriateness when working to eliminate health disparities, beyond a person’s primary language(51). Investigating the relevance of different constructs for specific groups, and how they are understood and interpreted, is vital to ensure adequate measures are being used in disaster research with minoritized populations (38,43). There is an obvious need for instrument development and assessment to eliminate data gaps and data collection most relevant to communities of color as those most at risk to climate change impacts. Valid instruments are needed to assess the disproportionate impacts of climate change on communities of color and inform climate equity interventions.

### Cognitive Mapping

Overall, our results revealed that participants cognitively considered disaster characteristics, resources, individual life facets, and felt effect when considering the survey items. In addition to this general cognitive process, individual disaster terms were often recalled within participants’ “think out loud” narratives about a specific disaster event, specific disaster location, or through the disaster’s unique antecedents and consequences. Since our themes and thematic mapping were developed using inductive reasoning from the interview data, many of our themes are novel and unique and not tied to existing literature. However, several of our findings corroborate theoretical and qualitative evidence among other fields and populations. Throughout the disaster planning and response professional fields, disasters are commonly defined and classified based on their onset, duration, effect, and recovery period. Thus, the disaster characteristics and felt effects themes reflect components of broadly shared mental paradigms among governments and professional disaster experts. Our resources theme is similar to the consideration of assets for stroke patients when contemplating disaster experiences and resilience(52). Specific life facets, such as experience or training as a military veteran, is known to have an impact on disaster perceptions and household disaster preparedness(22).

When mapping themes for the conceptualization of disaster/hazard/events (Figure 2), many expert panelists linked disaster characteristics (i.e., intensity, duration) with resources (i.e., personal). When answering a question about heating/ventilation/air conditioning failure, one participant noted, “I should quantify that…I have the income, this is not a disaster. In my mind a disaster is an event that supersedes available resources. I have enough…I am in a position that this is an inconvenience, it’s not a disaster.” This conceptualization is consistent with widely accepted views of disasters as events where the required capacity or ability to cope exceeds available resources(53). Another respondent stated, “Everyone is vulnerable depending on where you are, what you’re doing and what the impact is but there are certain people who are much more vulnerable because they don’t have the resources or the ability to be resilient.” In a recent multidisciplinary review of literature and concept analysis of household emergency preparedness, authors examined antecedents, or things that must occur prior to the concept (disaster) occurrence(54). Personal resources/financial capital was not included as an antecedent to household emergency preparedness. This may reflect a potential disconnect among USA-based disaster preparedness conceptualization and/or literature, and illustrates the importance of incorporating racial wealth disparities into the conversation of disaster-related disparities and climate resilience needs (58).

Past research has elucidated qualitative themes from rare disaster experiences, such as active shooter, within single event contexts (55). Similarly, even more common lived disaster experiences, such as hurricanes, are frequently researched relative to a single catastrophe(56). By sampling an expert panel, our study uniquely uncovered themes across multiple disaster experiences, settings, and events. As the incidence and prevalence of disaster compounding increases, understanding the relationships of disaster/hazard compounding and cascading will only become more relevant (57). Further research is warranted into cognitive mapping and participant understanding of specific disaster terms and categories as we presented here with unique mental schemas and cognitive processes relative to water contamination, hurricane, and active shooter that were unbound by any one specific disaster or event context. This may further elucidate how individuals affected make meaning, store and retrieve the memories of specific disaster events and event subtypes, which will inform additional vulnerability assessments and considerations needed beyond a general all-hazard approach.

### Linked Disaster Terms

We found a total of 379 unique instances of linked terms across the 18 interviews and 60 terms. Consideration of linked, co-occurring, and compounding disasters are especially relevant for conceptualizing the hazard vulnerability for those at higher risk for disaster consequences, such as those with Black or African American racial identities, older adults, and those with chronic respiratory disease(58). Disaster compounding has been observed at an increasing rate over the past decade, where one disaster event precipitates another, at times resulting in compounded, catastrophic losses(57). For example, a natural hazard like an earthquake could trigger a cascade of technological hazards, such as natural gas and power failure, water disruption, structural fires, and building or bridge collapses. Natural hazards and disasters compound the health impacts of other cascading or concurrent non-climate related disasters. In another example, Keith et al. (2021) noted how the hottest summer on record in the Northern Hemisphere coincided with the pandemic, requiring shifts in response resources and mitigation of compounding health hazards like heat-related injury and illness(59). Metzl et al. (2021) noted that pandemic-related resource shifting of first responders and emergency departments in major U.S. cities lead to increased lethality following recent multiple-victim shootings(60). Our findings reveal the need to further address and develop compounding and cascading risk considerations with a focus on events experienced or identified by a greater proportion those with Black or African American identities, such as the following elucidated from our pattern coding: water dam failure, evacuation, external flood, suspicious package/substance, and transportation failure. Climate resilience intersects with pre-existing health disparity, socio-economic wealth, systemic and structural racism in complex and interlaced ways. For example, the unmitigated history of maintenance, upkeep, and structural integrity of infrastructure such as water dams, levees, and transportation in communities of color diminishes climate resilience and increases the risk for compounding racial disparities in disaster effects and damage related to flooding, transportation, and evacuation(25,26).

### Transferability

When considering transferability, the findings reported here present a cognitive map and schema from an expert perspective relevant to all-hazards experiences, as well as those specific to disaster sub-types. These themes may be considered as an initial theory of a cognitive process map when considering individual and household vulnerability. We also identified several problematic hazard, disaster, and event terms that might not be interpreted by survey-takers as intended. Other measurements utilizing terms such as trauma, radiation exposure, or criminal acts of intent may require further validity testing and refinement in climate change research and practice. We further identified several hazard, disaster, or event terms that require clarification, elimination, replacement, or further revision in our validation process and survey instrument development. These are common terms, often used in organizational hazard vulnerability assessments, that may have relevance to other disaster and climate change research teams seeking to focus their work on individual or household units of analysis.

### Future Research

Here, we focused on household hazard vulnerability analysis instrument development with unique considerations for the problem of potential for data racial bias among those with Black or African American identities in the United States. This is an initial report in a multi-faceted process of survey development and validation, which includes additional content validity indices and validation with the intended population of older adults with chronic respiratory disease. We recommend our study be replicated among other populations who are at risk for the negative health impacts of climate related disasters, such as Indigenous people or those whose native language differs from the language in which their government’s business is conducted. The intersectionality of immigration and racial disparity for current generations of Black immigrants may present unique disaster risks that warrant further research. The questionnaire, themes, and information presented were generated from a predominantly Western-trained and educated panel. We suggest further validating the questions and themes presented in this study, to better fit regional and cultural nuances regarding the usefulness of the questions in other settings and contexts where cultural characteristics and relaying of information may differ from the Western lens with respect to disaster preparedness both domestically in the USA and abroad.

## Limitations

The findings from this study should be interpreted in light of the limitations of the design. We conducted the interviews during the COVID-19 pandemic, which elicited strong, compounding and consistent responses to items related to pandemic, epidemic, and infectious disease outbreaks that may not be as profound or present in the cognitive processes of future participants, outside the immediate pandemic context. The interviewer originated the videoconferences from a location with known contemporaneous drinking water contamination (Hawai’i, region impacted by Red Hill fuel facility), which may have influenced the priority, order, and recall primacy of thoughts related to water contamination for participants. The methodological contributions of the cognitive interviewing are to address the construct validity of survey items, and are not meant to generate inferential conclusions that generalize their responses to the broader population. Additional methods to address construct and other forms of validity are also required in instrument development and validation. We utilized an expert panel with extensive knowledge and experience in hazard and disaster situations both in the US and overseas. Given most of the disasters, hazards, or events were experienced by at least one of our expert panel members, this provides foundational knowledge that is more broadly transferable to people who have experienced disasters. However, additional validity and item testing is needed among the intended survey-takers, namely older adults with COPD to produce a valid survey instrument specifically for this population.

## Conclusion

This manuscript presents the findings of our initial cognitive interviews with an expert panel, conducted as the first step in the development of a household-level Hazard Vulnerability Analysis. Our novel methods were developed in order to detect and combat data racial bias in the instrument development specific to people with African American/Black racial identities. The findings reported here identified problematic hazard, disaster, and event terms that might not be interpreted by survey-takers as intended. We also identified specific items and terms that warrant further investigation as potentially identifying racial disparities in experiences or in cognitive interpretations. These findings informed our ongoing instrument development and revisions. Following the subsequent phases in this project, the instrument is being developed for patient-reported and clinician use to quantify risk and prioritize affirmative disaster preparedness interventions for those most vulnerable to climate-sensitive health risks and other disasters. This work informs precision public health directed at household disaster preparedness interventions, which is profoundly timely and important in the face of global climate change.

Funding: Research reported in this publication was supported by the National Institute on Minority Health And Health Disparities of the National Institutes of Health under Award Number R43MD017188 (PI: Castner). The content is solely the responsibility of the authors and does not necessarily represent the official views of the National Institutes of Health.

## CRediT author statement - <u>CRediT author statement</u> (elsevier.com)

**Taryn Amberson**: Investigation, Writing - Original draft preparation, visualization, supervision, project administration; **Olive Ndayishimiye**: Investigation, Initial Data Analysis, Resources, Writing - Review and editing^;^ **Quanah Yellow Cloud**: Investigation, Initial Data Analysis, Resources, Writing - Original draft preparation; **Jessica Castner**: Conceptualization, methodology, formal analysis, visualization, supervision, funding acquisition

## Researcher Characteristics

Researcher characteristics are transparently reported for readers to evaluate the potential for influence in the research, relationship with participants, and interactions in the qualitative interview and data analysis process. Our diverse team included those with biracial, Native American (Oglala Lakota), White, and Modern African diaspora identities. Our team included at least one member who identifies with the LGBTQ+ community. At the time of this study, the interviewer (female, White) held a Master of Public Health and a Bachelor of Science in Nursing degrees with board certifications as an emergency nursing and disaster healthcare professional. The same interviewer met with all twenty expert panelists.

## Data Availability

All data produced in the present study are available upon reasonable request to the authors.

## Appendix A Additional Detail for Methods

### Qualitative approach and research paradigm

The guiding theoretical framework for this work was Tourangeu’s analytic model of the survey response process (1). This model divides the survey response process into four stages: 1) comprehension of the question, 2) retrieval of relevant information from memory, 3) judgment and estimation process of the retrieved information, and 4) response processes (2).

Retrospective follow up probes, such as “What were you thinking when you answered this question?” or, “What led you to answer the way that you did?” were used to help researchers understand participants’ interpretative processes and help uncover interpretive and elusive errors through textual verification.

#### Sampling strategy

Our strategy intentionally over-sampled (up to 50%) those with Black and/or African American racial, biracial or multiracial identities as a novel strategy to combat racial data bias and develop a survey based on the population with the greatest demonstrated disparity for disaster-related assessment outcomes at the household level. (3, 4) Cognitive interviewing is generally conducted with 10 participants(2). We intentionally doubled this sample size in order to include a full cognitive interviewing sample size panel of those with Black and/or African American racial identities. These factors, along with the vast geographical variability of hazards/disasters, were considered when determining a sample size of 20 respondents.

#### Data Processing and Analysis

The cognitive interviewing portion of the first 18 interviews consisted of: 1) questions about Experts’ personal and professional experiences with 60 different types of hazards/disaster, and 2) Demographics. Using standard cognitive interviewing methodologies, participants were asked to think out loud to tell the interviewer the story of why they answered the way that they did. The interviewer actively considered participant comprehension, recall, judgment, and response and applied standard probes where more information was needed to clarify the participant’s cognitive processes. The final two respondents were interviewed using a finalized instrument draft to assess the need for additional reparative procedures. The survey was modified in two rounds of content validity indices not covered in this initial manuscript and to be reported elsewhere.

We utilized secure project Google docs, Google Sheets, and Q Notes as software to complete these analytic steps. Q-Notes is a publicly available software application maintained by National Center for Health Statistics designed specifically for cognitive interview methods(5).

Using the audio-visual recorded interview as the data source, analysis was conducted as follows: key text summaries were extracted for each respondent about how they interpreted the hazard/disaster term and generated an answer. This within-participant analytic step includes ascertaining the participant’s explanation of their thinking process and problems the participant experienced with the question-answer process. Next, we synthesized these textual terms and summaries across all respondents for each hazard/disaster term to inductively generate and map emerging thematic codes. These theme codes were visualized into a figure as a schema as tree branches across participants and for each disaster/hazard term. Pattern coding was used to compare the responses (yes/no) to ever experiencing the hazard/disaster in the participant’s lifetime. The pattern coding was utilized to ascertain differences between those who identify their race as B/AA (inclusive of biracial/multi-racial) and those who did not identify their race as B/AA at the time of the interview. We utilized the analytic findings from each of the previous steps listed here to draw conclusions about the performance of each item across all participants and elucidate potential for racial data bias in the content validity performance of the individual items.

## Appendix B - Cognitive Interview Item Example Household Disaster Hazard Vulnerability

**<u>1. Active Shooter</u>**

**AS_LI2. In your lifetime, have you ever been directly impacted by?: Active Shooter**

_ Yes

_ No

## Appendix A References

(1) Miller K, Chepp V, Willson S, Padilla J. Cognitive interviewing methodology. : John Wiley & Sons; 2014.

(2) Willis GB. Cognitive Interviewing Revisited: A Useful Technique, in Theory? Methods for Testing and Evaluating Survey Questionnaires Hoboken, NJ, USA: John Wiley & Sons, Inc; 2004. p. 23–43.

(3) McQuade L, Merriman B, Lyford M, Nadler B, Desai S, Miller R, et al. Emergency Department and Inpatient Health Care Services Utilization by the Elderly Population: Hurricane Sandy in The State of New Jersey. Disaster medicine and public health preparedness 2018 Dec;12(6):730–738.

(4) Flores AB, Collins TW, Grineski SE, Chakraborty J. Disparities in Health Effects and Access to Health Care Among Houston Area Residents After Hurricane Harvey. Public health reports (1974) 2020 Jul;135(4):511–523.

(5) Collaborating Center for Questionnaire Design and Evaluation Research. Cognitive Interviewing: Data Collection and Interviewing Procedures. 2022; Available at: https://www.n.cdc.gov/QNotes/Project/Report. Accessed July 29, 2022.

## References

(1) World Health Organization. Climate change and health. 2021; Available at: https://www.who.int/news-room/fact-sheets/detail/climate-change-and-health. Accessed June 14, 2022.

(2) United States Geological Survey. How can climate change affect natural disasters? n.d.; Available at: https://www.usgs.gov/faqs/how-can-climate-change-affect-natural-disasters#:~:text=With%20increasing%20global%20surface%20temperatures,more%20powerful%20storms%20to%20develop. Accessed June 14, 2022.

(3) Gotanda H, Fogel J, Husk G, Levine JM, Peterson M, Baumlin K, et al. Hurricane Sandy: Impact on Emergency Department and Hospital Utilization by Older Adults in Lower Manhattan, New York (USA). Prehospital and disaster medicine 2015 Oct;30(5):496–502.

(4) Quast T. Emergency Department Visits by and Hospitalizations of Senior Diabetics in the Three Years Following Hurricanes Katrina and Rita. EconDisCliCha 2019 Jan 15,;3(2):151–160.

(5) NOAA National Centers for Environmental Information (NCEI). U.S. Billion-Dollar Weather and Climate Disasters. 2022; Available at: https://www.ncdc.noaa.gov/billions/. Accessed February 19, 2020.

(6) Centre for Research on the Epidemiology of Disasters. 2021 Disasters in numbers. 2022; Available at: https://cred.be/sites/default/files/2021_EMDAT_report.pdf. Accessed June 14, 2022.

(7) Assistant Seretary for Preparedness and Response, U.S. Department of Health and Human Services. Hazard vulnerability/risk assessment. 2022; Available at: https://asprtracie.hhs.gov/technical-resources/3/hazard-vulnerability-risk-assessment/1. Accessed June 14, 2022.

(8) Campbell P, Trockman SJ, Walker AR. STRENGTHENING HAZARD VULNERABILITY ANALYSIS: RESULTS OF RECENT RESEARCH IN MAINE. Public health reports (1974) 2011 Mar 1,;126(2):290–293.

(9) California Hospital Association. Hazard vulnerability analysis. 2021; Available at: https://www.calhospitalprepare.org/hazard-vulnerability-analysis. Accessed June 14, 2022.

(10) Lieberman-Cribbin W, Liu B, Sheffield P, Schwartz R, Taioli E. Socioeconomic disparities in incidents at toxic sites during Hurricane Harvey. J Expo Sci Environ Epidemiol 2021 Apr 19.

(11) Lawrence WR, Lin Z, Lipton EA, Birkhead G, Primeau M, Dong G, et al. After the Storm: Short-term and Long-term Health Effects Following Superstorm Sandy among the Elderly. Disaster medicine and public health preparedness 2019 Feb;13(1):28–32.

(12) McQuade L, Merriman B, Lyford M, Nadler B, Desai S, Miller R, et al. Emergency Department and Inpatient Health Care Services Utilization by the Elderly Population: Hurricane Sandy in The State of New Jersey. Disaster medicine and public health preparedness 2018 Dec;12(6):730–738.

(13) Sirey JA, Berman J, Halkett A, Giunta N, Kerrigan J, Raeifar E, et al. Storm Impact and Depression Among Older Adults Living in Hurricane Sandy-Affected Areas. Disaster medicine and public health preparedness 2017 Feb;11(1):97–109.

(14) Swerdel JN, Rhoads GG, Cosgrove NM, Kostis JB. Rates of Hospitalization for Dehydration Following Hurricane Sandy in New Jersey. Disaster medicine and public health preparedness 2016 Apr;10(2):188–192.

(15) Shih RA, Acosta JD, Chen EK, Carbone EG, Xenakis L, Adamson DM, et al. Improving Disaster Resilience Among Older Adults: Insights from Public Health Departments and Aging-in-Place Efforts. Rand health quarterly 2018 Aug;8(1):3.

(16) Wyte-Lake T, Claver M, Dobalian A. Assessing Patients’ Disaster Preparedness in Home-Based Primary Care. Gerontology (Basel) 2016 Apr;62(3):263–274.

(17) Wyte-Lake T, Claver M, Griffin A, Dobalian A. The Role of the Home-Based Provider in Disaster Preparedness of a Vulnerable Population. Gerontology (Basel) 2014 Jun;60(4):336–345.

(18) Casey-Lockyer M, Heick RJ, Mertzlufft CE, Yard EE, Wolkin AF, Noe RS, et al. Deaths Associated with Hurricane Sandy — October–November 2012. MMWR. Morbidity and Mortality Weekly Report 2013 May 24,;62(20):393–397.

(19) Bell SA, Donnelly JP, Li W, Davis MA. Hospitalizations for chronic conditions following hurricanes among older adults: A self?controlled case series analysis. Journal of the American Geriatrics Society (JAGS) 2022 Jun;70(6):1695–1703.

(20) Ko JY, Strine TW, Allweiss P. Chronic Conditions and Household Preparedness for Public Health Emergencies: Behavioral Risk Factor Surveillance System, 2006-2010. Prehospital and disaster medicine 2014 Feb;29(1):13–20.

(21) DeBastiani SD, Strine TW, Vagi SJ, Barnett DJ, Kahn EB. Preparedness Perceptions, Sociodemographic Characteristics, and Level of Household Preparedness for Public Health Emergencies: Behavioral Risk Factor Surveillance System, 2006-2010. Health security 2015 Sep 1,;13(5):317–326.

(22) Der-Martirosian C, Strine T, Atia M, Chu K, Mitchell MN, Dobalian A. General Household Emergency Preparedness: A Comparison Between Veterans and Nonveterans. Prehospital and disaster medicine 2014 Apr;29(2):134–140.

(23) Arias E, Tejada-Vera B, Ahmad F, Kochanek KD. Provisional life expectancy estimates for 2020. National Center for Health Statistics 2021 Jul.

(24) Chidambaram P, Neuman T, Garfield R. Racial and Ethnic Disparities in COVID-19 Cases and Deaths in Nursing Homes. 2020 Oct.

(25) National Academies of Sciences, Engineering, and Medicine. Equitable and Resilient Infrastructure Investments. Washington, DC: The National Academies Press; 2022.

(26) Aune KT, Gesch D, Smith GS. A spatial analysis of climate gentrification in Orleans Parish, Louisiana post-Hurricane Katrina. Environmental research 2020 Jun;185:109384.

(27) Shukla M, Gibson-Scipio WM, Amberson T, Castner J. Disaster Preparedness Disparities: Analysis of the 2018-2020 FEMA National Household Survey. American Journal of Respiratory and Critical Care Issues 2022;205:A3853.

(28) Weber L, Hilfinger Messias DK. Mississippi front-line recovery work after Hurricane Katrina: an analysis of the intersections of gender, race, and class in advocacy, power relations, and health. Soc Sci Med 2012 Jun;74(11):1833–1841.

(29) Gauthier GR, Smith JA, García C, Garcia MA, Thomas PA. PMC7454830; Exacerbating Inequalities: Social Networks, Racial/Ethnic Disparities, and the COVID-19 Pandemic in the United States. J Gerontol B Psychol Sci Soc Sci 2021 Feb 17;76(3):e88–e92.

(30) Davis JR, Wilson S, Brock-Martin A, Glover S, Svendsen ER. The impact of disasters on populations with health and health care disparities. Disaster medicine and public health preparedness 2010;4(1):30–38.

(31) Ali JS, Farrell AS, Alexander AC, Forde DR, Stockton M, Ward KD. PMC5411306; Race differences in depression vulnerability following Hurricane Katrina. Psychol Trauma 2017 May;9(3):317–324.

(32) Thomas DSK, Jang S, Scandlyn J. PMC7467012; The CHASMS conceptual model of cascading disasters and social vulnerability: The COVID-19 case example. Int J Disaster Risk Reduct 2020 Dec;51:101828.

(33) Alexander M. The New Jim Crow. La Vergne: The New Press; 2020.

(34) Rooks N. Cutting school: privatization, segregation, and the end of public education. : New Press; 2017.

(35) Taylor K. Race for profit. Chapel Hill: University of North Carolina Press; 2019.

(36) Castner J, Barnett R, Moskos LH, Folz RJ, Polivka B. Home environment allergen exposure scale in older adult cohort with asthma. Can J Public Health 2020 Jun 16,;112(1):97–106.

(37) Castner J. Precision Assessment Algorithm for Reducing Disaster-related Respiratory Health Disparities. 2021; Available at: https://reporter.nih.gov/search/KqjFrLCsPUmiZFuqr_DE2A/project-details/10401726. Accessed August 18, 2022.

(38) Miller K, Chepp V, Willson S, Padilla J. Cognitive interviewing methodology. : John Wiley & Sons; 2014.

(39) Collaborating Center for Questionnaire Design and Evaluation Research. Cognitive Interviewing: Data Collection and Interviewing Procedures. 2022; Available at: https://wwwn.cdc.gov/QNotes/Project/Report. Accessed July 29, 2022.

(40) US Department of Health and Human Services, Assistant Secretary for Preparedness and Response. Risk Identification and Site Criticality (RISC) Toolkit. 2019; Available at: https://www.hsdl.org/c/risc-toolkit/. Accessed August 10, 2022.

(41) Centre for Research on the Epidemiology of Disasters. General Classification. 2009;. Accessed July 18, 2022.

(42) Willis GB. Cognitive Interviewing Revisited: A Useful Technique, in Theory? Methods for Testing and Evaluating Survey Questionnaires Hoboken, NJ, USA: John Wiley & Sons, Inc; 2004. p. 23–43.

(43) Willis GB. Cognitive interviewing: A tool for improving questionnaire design. Thousand Oaks [u.a.]: Sage; 2004.

(44) Willis GB. Analysis of the Cognitive Interview in Questionnaire Design. Cary: Oxford University Press, Incorporated; 2015.

(45) Centers for Disease Control and Prevention, Collaborating Center for Questionnaire Design and Evaluation Research. Q-Bank: Improving Surveys by Sharing Knowledge. 2021; Available at: https://wwwn.cdc.gov/qbank/Search/Reports.aspx#/Reports. Accessed August 18, 2022.

(46) Heagele TN, Adams LM, McNeill CC, Alfred DM. Validation and Revision of the Household Emergency Preparedness Instrument (HEPI) by a Pilot Study in the City University of New York. Disaster medicine and public health preparedness 2022 Mar 25,:1–9.

(47) Centers for Disease Control and Prevention. Community Assessment for Public Health Emergency Response (CASPER) Toolkit: 3rd Edition. 2019.

(48) Couig MP, Travers JL, Polivka B, Castner J, Veenema TG, Stokes L, et al. At-Risk populations and public health emergency preparedness in the United States: Nursing leadership in communities. Nursing outlook 2021 Jul;69(4):699–703.

(49) Mailloux NA, Henegan CP, Lsoto D, Patterson KP, West PC, Foley JA, et al. Climate Solutions Double as Health Interventions. International journal of environmental research and public health 2021 Dec 18,;18(24):13339.

(50) Berberian AG, Gonzalez DJX, Cushing LJ. Racial Disparities in Climate Change-Related Health Effects in the United States. Curr Envir Health Rpt 2022 May 28,;9(3):451–464.

(51) Burlew AK, Ph.D, Feaster D, Ph.D, Brecht M, Ph.D, Hubbard, Robert, Ph.D., M.B.A. Measurement and data analysis in research addressing health disparities in substance abuse. Journal of substance abuse treatment 2009;36(1):25–43.

(52) O’Sullivan TL, Fahim C, Gagnon E. Asset Literacy Following Stroke: Implications for Disaster Resilience. Disaster medicine and public health preparedness 2018 Jun;12(3):312–320.

(53) United Nations Office for Disaster Risk Reduction. Disaster. n.d.; Available at: https://www.undrr.org/terminology/disaster. Accessed August 20, 2022.

(54) Wilcox L, Heagele T, McNeill C. Household emergency preparedness: A multidisciplinary concept analysis. Nursing forum (Hillsdale) 2022 Mar;57(2):305–310.

(55) McCall WT. Caring for Patients From a School Shooting: A Qualitative Case Series in Emergency Nursing. Journal of emergency nursing 2020 Sep;46(5):712-721.e1.

(56) Adams V, Kaufman SR, van Hattum T, Moody S. Aging Disaster: Mortality, Vulnerability, and Long-Term Recovery among Katrina Survivors. Medical anthropology 2011 May 1,;30(3):247–270.

(57) Liu, Minquan and Huang, Michael. Compound Disasters and Compounding Processes. The United Nations Office for Disaster Risk Reduction 2014 Jan 8,.

(58) National Academies of Sciences, Engineering, and Medicine. Perspectives on Climate and Environmental Justice on the U.S. Gulf Coast: Proceedings of a Webinar—in Brief. 2021.

(59) Keith L, Iroz-Elardo N, Austof E, Sami I, Arora M. Extreme heat at outdoor COVID-19 vaccination sites. The journal of climate change and health 2021 Oct;4:100043.

(60) Metzl JM, Piemonte J, McKay T. Mental Illness, Mass Shootings, and the Future of Psychiatric Research into American Gun Violence. Harvard review of psychiatry 2021 Jan;29(1):81–89.

